# Biologic correlates of beneficial convalescent plasma therapy in a COVID-19 patient reveal disease resolution mechanisms

**DOI:** 10.1101/2022.02.03.22269612

**Authors:** Natalie Bruiners, Valentina Guerrini, Rahul Ukey, Ryan Dikdan, Jason Yang, Pankaj Kumar Mishra, Alberta Onyuka, Deborah Handler, Joshua Vieth, Mary Carayannopulos, Shuang Guo, Maressa Pollen, Abraham Pinter, Sanjay Tyagi, Daniel Feingold, Claire Philipp, Steven Libutti, Maria Laura Gennaro

## Abstract

**Background:** While the biomarkers of COVID-19 severity have been thoroughly investigated, the key biological dynamics associated with COVID-19 resolution are still insufficiently understood.

**Main body:** We report a case of full resolution of severe COVID-19 due to convalescent plasma transfusion in a patient with underlying multiple autoimmune syndrome. Following transfusion, the patient showed fever remission, improved respiratory status, and rapidly decreased viral burden in respiratory fluids and SARS-CoV-2 RNAemia. Longitudinal unbiased proteomic analysis of plasma and single-cell transcriptomics of peripheral blood cells conducted prior to and at multiple times after convalescent plasma transfusion identified the key biological processes associated with the transition from severe disease to disease-free state. These included (i) temporally ordered upward and downward changes in plasma proteins reestablishing homeostasis and (ii) post-transfusion disappearance of a particular subset of dysfunctional monocytes characterized by hyperactivated Interferon responses and decreased TNF-α signaling.

**Conclusions:** Monitoring specific subsets of innate immune cells in peripheral blood may provide prognostic keys in severe COVID-19. Moreover, understanding disease resolution at the molecular and cellular level should contribute to identify targets of therapeutic interventions against severe COVID-19.

## Introduction

Almost two years into the COVID-19 pandemic caused by the severe acute respiratory syndrome coronavirus 2 (SARS-CoV-2), we still have a limited therapeutic armamentarium to prevent or treat the severe outcomes of this disease (reviewed in ^1^). Current therapeutic recommendations for COVID-19 by the US Food and Drug Administration (FDA) include antivirals such as remdesivir and anti-inflammatory drugs such as corticosteroids for hospitalized patients requiring supplemental oxygen, and monoclonal antibodies directed against the SAS-CoV-2 Spike protein for outpatients at high risk of disease progression ^2^. New oral antiviral drugs have received FDA emergency use authorization (EUA) for vulnerable individuals to reduce the risk of COVID-19-related hospitalization ^3,4^. However, additional analyses have shown efficacy levels for these drugs well below those detected in the interim analyses ^5^. The recently emerged Omicron variant may constitute a new challenge for monoclonal antibody therapy, since mutations in key epitopes may reduce variant susceptibility to the therapy ^6^. Convalescent plasma therapy received EUA from the FDA to treat COVID-19 in hospitalized patients in March 2021 (https://www.fda.gov/media/141477/download). This EUA was based on precedents of passive immunotherapy for various respiratory infectious diseases^7^, and on preclinical evidence on safety and efficacy. However, the effectiveness of passive immunotherapy remains controversial. Observational studies have shown that plasma therapy reduces mortality, particularly when administered early during hospitalization ^8-10^, consistent with favorable clinical outcomes when seroconversion and production of neutralizing antibodies occur early in infection ^11^. Upon review of 16 randomized controlled trials including >16,000 patients, the World Health Organization has recently deliberated that convalescent plasma therapy shows no benefits in non-severe COVID-19, but that clinical trials for its use in severe and critical COVID-19 are still warranted ^12^. Thus, whether convalescent plasma therapy can be outcome-determining, how, and in which kind of patients, remains an open question.

Key elements of potentially successful plasma therapy for COVID-19 are understudied. First, little is known about indicators of the beneficial effects of convalescent plasma therapy in the recipient’s plasma. Most observational studies and clinical trials have only reported on routine clinical assessments (for example ^8,13^) and, in a few cases, on coagulation markers and few proinflammatory cytokines ^9,14^. Second, it can be inferred that plasma therapy may benefit COVID-19 patients having compromised humoral immunity or receiving B-cell depleting therapy for multiple sclerosis, rheumatic and autoimmune diseases, or hematologic malignancies ^15-17^. Indeed, beneficial effects of passive immunotherapy on clinical course have been documented, for example, for a patient with humoral immunodeficiency ^18^. That report focused on the properties of donor plasma, importantly highlighting how the assessment of convalescent plasma therapy effects should require knowledge on key characteristics of the donor plasma, such as its neutralizing capacity. However, the report did not investigate viral load changes and host correlates of COVID-19 resolution in the plasma recipient. Such knowledge would help identify the biological mechanisms associated with the beneficial effects of convalescent plasma therapy and, more generally, with the resolution of severe COVID-19. Here, in addition to characterizing key donor plasma properties, we report the in-depth analysis of viral and host biologic correlates of the clinical resolution of COVID-19 by convalescent plasma therapy in a patient with underlying immunological disorders.

## Results

### Clinical course and transfusion effects on viral load and clinical parameters

In March 2020, a Caucasian woman in her fifties with multiple autoimmune syndrome (MAS) developed flu-like symptoms (Figure 1A). Her treatments for MAS included daily hydroxychloroquine, weekly methotrexate, and every 5-6 months Rituximab infusion. Three days after symptom onset, she presented to the emergency department (ED), where the nasopharyngeal fluid PCR test was positive for SARS-CoV-2 and chest X ray (CXR) examination demonstrated early right lower lung infiltrates. She was discharged with Azithromycin dose pack. On day 14, she returned to the ED with fever (101.2°F) and hypoxia (pulse oximetry of 87% on ambient air). CXR showed bilateral lung opacities with interval progression from the previous exam. She was placed on 4-liter supplemental oxygen and admitted to a hospital. On day 16, she was started on a 10-day remdesivir course. The hospital course was complicated by hypotension (68/44 mm Hg) and hypothermia (93.1°F) (Figure 1B), requiring transfer to the medical intensive care unit (MICU) and pharmacologic blood pressure support.

**Figure 1.**
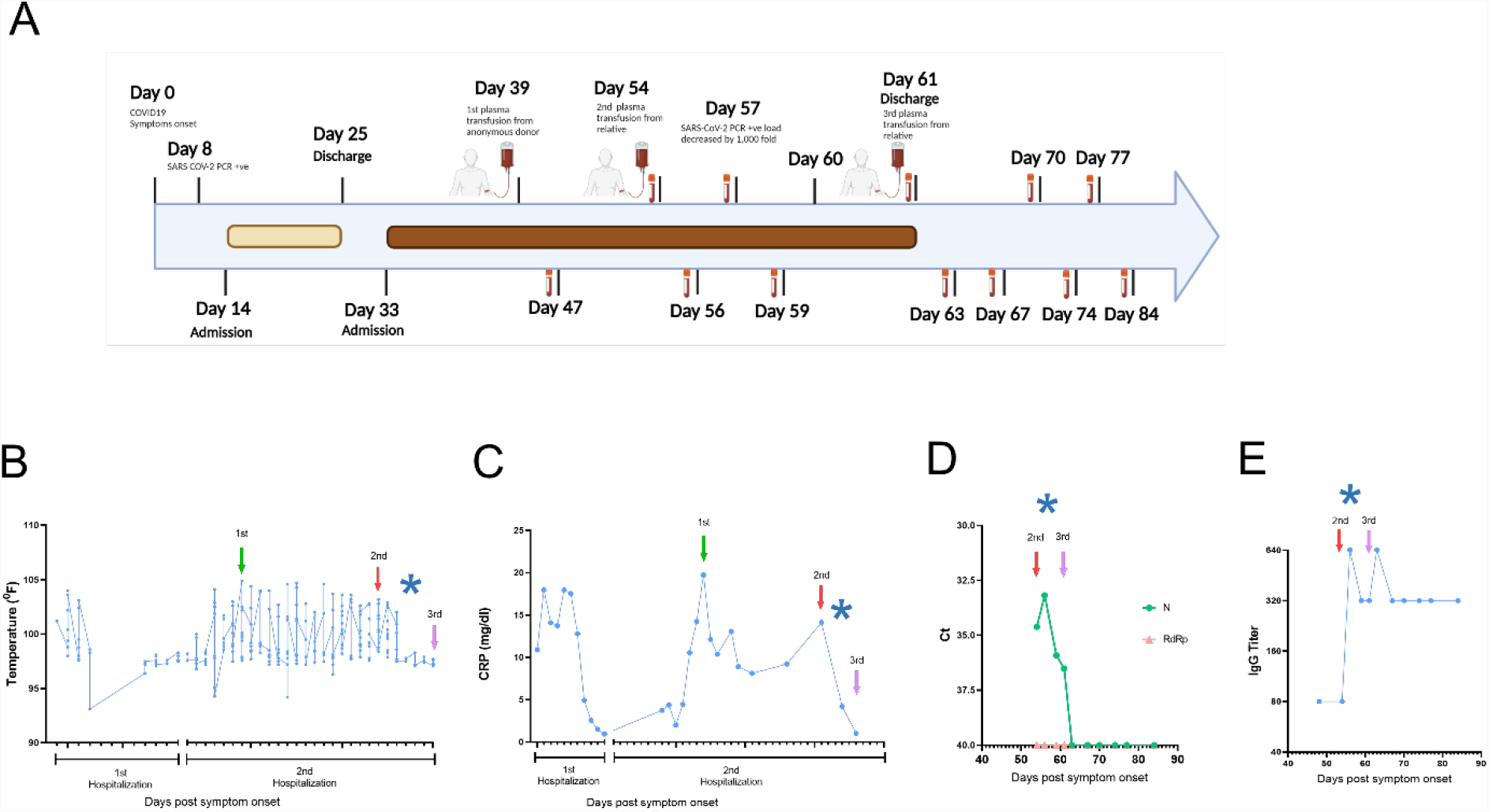
Timeline of the clinical course. **(A)** Day 0 indicates the date of the COVID-19 symptom onset. The days post-onset at which COVID-19 related symptoms, hospitalization course, RT-PCR test results, convalescent plasma transfusions, and interventions took place are indicated. **(B-E)** Longitudinal analysis of **(B)** maximum body temperature, **(C)** C-Reactive protein (CRP) in plasma, (**D)** SARS-CoV-2 RNA detection in plasma by qRT-PCR, **(E)** Titers for IgG against SARS-CoV-2 Spike Receptor binding domain (RBD). The y axis in each panel indicates the corresponding measurement unit. In all panels, the x axis indicates the day numbering as depicted in panel A. In panels B-C, the timeline in the x axis highlights the first hospitalization (days 14-25) and the second hospitalization (days 33-61), since the corresponding measurements were performed only in the hospital. In all panels, the vertical arrows indicate convalescent plasma transfusions from anonymous donor (1^st^) and from a patient’s relative (2^nd^ and 3^rd^). The asterisk indicates the drop in viral load recorded at day 57 (in panel A). In panel D, Ct, cycle threshold; N, SARS-CoV-2 nucleocapsid gene; RdRP, SARS-CoV-2 RNA-dependent RNA polymerase gene.

On day 22, a computed tomography (CT) angiogram of the chest showed bilateral COVID-19 pneumonia and a segmental artery pulmonary embolism; anticoagulation was increased from prophylactic to therapeutic dosing. Fevers resolved, and the patient was weaned off of supplemental oxygen at rest and discharged after completing 10-day course of remdesivir. On day 33, she was re-admitted to the hospital with a fever of 102°F and pulse oximetry of 93% on ambient air. She was started empirically on supplemental oxygen via nasal cannula, piperacillin/tazobactam, vancomycin, and prednisone. She continued to have high fevers twice daily, accompanied by increasing C-reactive protein (CRP) levels (Figure 1B and C). The patient received 400 mL of COVID-19 convalescent plasma from an anonymous donor on day 40; however, fevers persisted daily reaching maximum temperature of 104.7°F and CT of the chest with contrast on day 45 showed worsened bilateral ground-glass lung opacities compared to one week prior. She continued to require supplemental oxygen via nasal cannula (2 liters). Clinical flow cytometry tests detected no B cells in the patient’s peripheral blood (data not shown), presumably due to the use of rituximab and/or methotrexate, since treatment with these drugs can reduce B cell frequencies and humoral immune responses (for example ^19,20^). On day 48, plasma samples from the patient and family members were analyzed in our research laboratory for antibody testing. The patient exhibited undetectable IgG titers (not shown), while the patient’s relative, who had recovered from COVID-19, had high anti-SARS-CoV-2 Receptor binding domain (RBD) IgG titers (> 1:5,120) (Figure S1A), mostly of the IgG1 subclass (Figure S1B). Further characterization of the relative’s plasma showed high neutralizing activity against live SARS-CoV-2 [neutralizing titer 50 (NT50) = 1:160] (Figure S1C), which resided in the RBD-specific antibodies, since the neutralizing activity of the plasma was abrogated when plasma samples were depleted from anti-RBD antibodies but not from antibodies against a control SARS-CoV-2 antigen (nucleocapsid) (Figure S1D). On day 54, the patient received 600 mL of her relative’s convalescent plasma. Oxygen saturation improved and patient no longer required supplemental oxygen at rest on day 54. On day 57, SARS-CoV-2 PCR analysis of a nasopharyngeal sample showed a ∼1000-fold decrease [10 cycle threshold (Ct) values] in viral burden relative to the preceding test result. Concurrently, fevers abruptly resolved on day 57 (Figure 1B) and plasma markers of inflammation, such as C reactive protein (CRP), decreased rapidly (Figure 1C). SARS-CoV-2 PCR testing of a nasopharyngeal sample became negative on day 59 and lowly positive on day 60. The patient remained afebrile. Oxygen saturation remained normal at rest. She received a second transfusion of 400 mL of her relative’s convalescent plasma on day 61 and was discharged from the hospital. The SARS-CoV-2 PCR test remained stably negative after this transfusion for >20 days (Figure 1A), after which testing was discontinued. Antibody binding assays performed for >20 days after her discharge showed stable levels of anti-RBD IgG (Figure 1D). Peripheral blood was collected from the patient at multiple times pre- and post-transfusion with her relative’s plasma for downstream analyses, as indicated in Figure 1A. We also measured the abundance of plasma viral RNA before and after this transfusion (day 54 through day 84 post symptom onset) utilizing oligonucleotide primers and probes for the nucleocapsid (N) and RNA-dependent RNA polymerase (RdRp) genes of SARS-CoV-2. Viral RNA was detected with the N-specific probes on day 54 through 61 and became undetectable from day 63 onward, following her second transfusion with the relative’s plasma (Figure 1E). Thus, the drop of viral RNA signal in plasma was delayed approximately two days relative to the signal drop in the nasopharyngeal fluid. No signal was detected for the RdRp gene across samples (Figure 1E). The signal discrepancy observed for the two viral genes strongly suggests that the viral RNA detected in plasma is not genomic but that it rather derives from viral transcripts in infected cells. This possibility is consistent with the N transcript being the most abundant viral RNA species in infected cells ^21^. SARS-CoV-2 RNAemia has been associated with reduced anti-SARS-CoV-2 IgG levels and neutralization capacity of plasma and with severe or fatal COVID-19 ^22,23^. Thus, administration of the relative’s convalescent plasma resulted in fever remission, improved respiratory status, return to normal of plasma indicators of inflammation, and negativization of plasma and upper respiratory tract fluids for viral RNA.

### Plasma cytokine and proteome self-organize temporally and identify key biological processes during disease resolution

Utilizing blood samples collected for research purposes, we performed cytokine and proteomic analysis of plasma collected from the patient collected from the patient on the day of (prior to) the first transfusion with her relative’s plasma (i.e., day 54 post-symptom onset), and multiple times thereafter (see Fig. 1A for blood collection points). Unbiased hierarchical clustering analysis and principal component analysis revealed dynamic changes in plasma cytokine and protein concentrations over four temporal phases following transfusion (Figure 2A-D). Changes in the abundance of plasma proteins became detectable between days 56 and 61 (Figure 2A-B) -- a few days after the transfusion, and slightly preceded those in plasma cytokines. Between days 61 and 67, the concentration of most plasma cytokines decreased sharply, showing that disease resolution was linked with countering the hyperproduction of inflammatory cytokines typically associated with severe COVID-19 ^24^. Plasma proteins exhibited a more complex behavior, with some proteins decreasing (Figure 2B, orange vertical arrow) and others increasing (Figure 2B, green vertical arrow) following transfusion. We call these two protein subsets Module A and Module B, respectively (see Figure S3 for contribution to Principal Component 1 of the two Modules).

**Figure 2.**
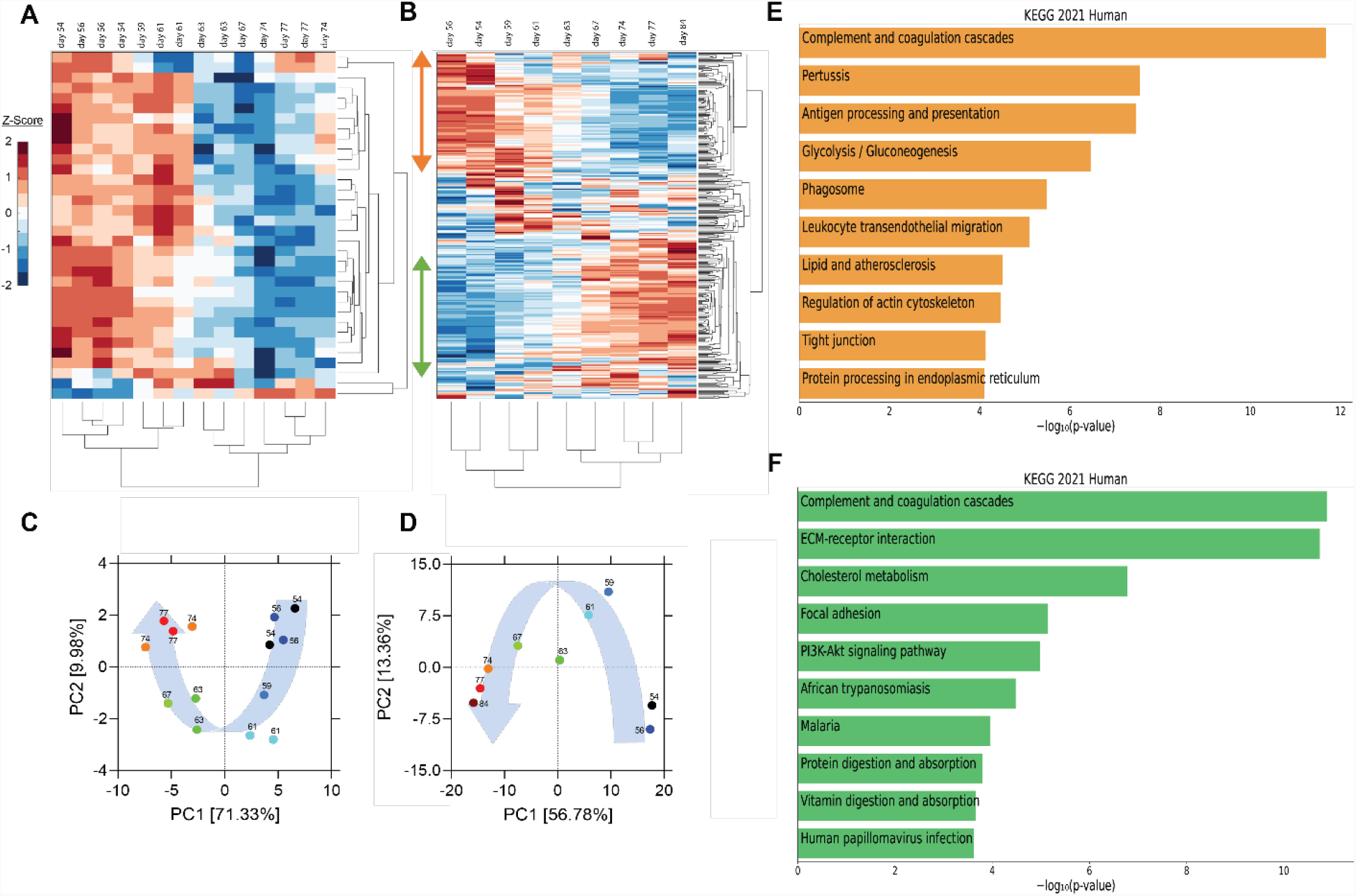
Unsupervised hierarchical clustering and principal component analysis (PCA) of cytokines and proteins in the recipient’s plasma before and after convalescent plasma transfusion. (A-B) Unsupervised hierarchical clustering was performed on cytokine profile and proteome using MATLAB. Vertical arrows in panel B mark the set of proteins that decrease (orange arrow) or increase (green arrow) following transfusion. (C-D) PCA was conducted on plasma cytokine profile (C) and proteome (D). (E-F) The top 10 enriched KEGG pathways for proteins that decrease (orange bars) (referred to as module A in Fig. S3 and in the text) or increase (green bars) (module B in Fig. S3 and in the text) using EnrichR Pathway Analysis.

By day 74 post-transfusion, plasma cytokines and proteins reached their steady-state concentrations, indicating therapeutic resolution. Bioinformatic analyses of plasma proteins in Modules A and B is presented below.

Pathway enrichment analysis using the Enrichr tool ^25^ identified “complement and coagulation cascades” as top term in both Module A and Module B (Figure 2EF), reflecting the known complement and coagulation disorders associated with severe disease by SARS-CoV-2 and other pathogenic coronaviruses ^26,27^. Proteins included in this term in Module A (Figure 2E) were predominantly components of the complement cascade (Table 1), consistent with complement hyperactivation potentially causing host tissue damage and disease ^28^ and being associated with severe COVID-19 ^29^. The second top term in Module A is “pertussis” (Figure 2E). In addition to proteins also present in “complement and coagulation cascades”, this pathway included cluster of differentiation 14 (CD14) (Table 1). Soluble CD14 (sCD14) and sCD163, which we also find decreased in the patient’s plasma following transfusion, contribute to monocyte-macrophage activation and increase with COVID-19 severity ^30^. Indeed, CD14 is currently being explored as a COVID-19 therapeutic target (clinical trial NCT04391309). The third pathway in Module A was “antigen processing and presentation” (Figure 2E). This pathway includes heat shock proteins (HSP) (Table 1) that have been described as “facilitators” of infections by several viruses, including coronaviruses and Zika ^31 32^. Some, such as HSP90, have been proposed as targets of therapeutic strategies against viral replication ^33,34^. The “antigen processing and presentation” pathway also included human leukocyte antigen (HLA) class I molecules and β2-microglobulin (β2-m), which is essential for the conformation of the major histocompatibility complex (MHC) class I protein complex (Table 1). Blood levels of soluble HLA antigens are elevated during chronic infection with hepatitis virus B and C and decrease following treatment of chronic viral infections ^35^, suggesting an association with viral persistence. Moreover, high β2-m levels in blood have been associated with several viral infections, including COVID-19 ^36,37^. An additional member of the “antigen processing and presentation” pathway is Calreticulin (Table 1), a molecular chaperone ensuring proper folding and functioning of proteins in the endoplasmic reticulum (ER) ^38^. Since all coronaviruses use the ER to replicate, severe infection with these viruses and the consequent massive production of viral proteins may induce ER stress and activate a compensatory mechanism known as the unfolded protein response, which can further enhance viral replication ^39^. Taken together, the analysis of the top three terms in Module A shows that clinical resolution of COVID-19 due to convalescent plasma transfusion was accompanied by the longitudinal decrease of plasma markers associated with pathogenic or harmful functions. These included (i) dampening of the excessive activation of the complement cascade and hyperinflammatory responses, with consequent reduction of their tissue- and organ-damaging effects, and (ii) decreased expression of antigen presentation and various protein modifying systems that may favor viral replication.

**Table 1.**
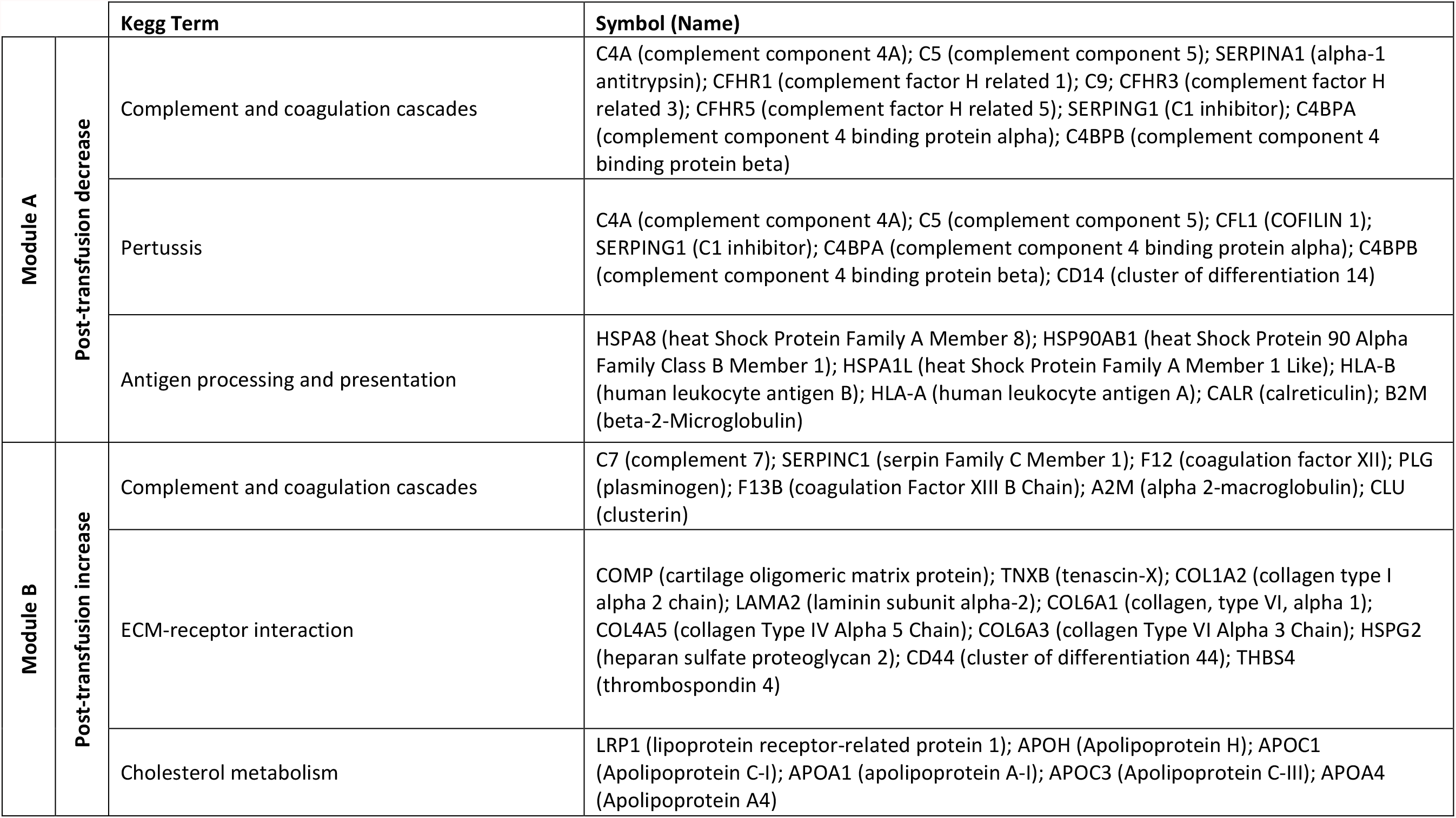
Proteins in the top three KEGG pathways that decrease (module A) or increase (module B) post-transfusion

As mentioned above, the top term in the pathway analysis in Module B, which increased with convalescent plasma transfusion, was also ‘complement and coagulation cascades’ (Figure 2F). Proteins assigned to this term in Module B included mostly coagulation factors that were low prior to the convalescent plasma transfusion, and serine protease inhibitors (Table 1). In addition, this pathway included components of the kallikrein-kinin system (Table 1), which participates in coagulation and control of blood pressure and may contribute to lung angioedema in severe COVID-19 ^40^. The second pathway in Module B is “Extracellular matrix (ECM) receptor interaction” (Figure 2F and Table 1). Interactions between ECM constituents and cells, which are typically mediated by transmembrane molecules, contribute to the regulation of key cellular functions, including adhesion, migration, and differentiation ^41^. Proteins associated with ECM receptors and focal adhesion (the fourth topmost pathway in Module B, Figure 2F) have been found decreased in the lung tissue of patients deceased with COVID-19 pneumonia, suggesting dysregulation of the lung extracellular microenvironment ^42^. The observed increased plasma levels of these proteins following convalescent plasma transfusion – if they reflect changes occurring also in organ microenvironments – could then be considered a meaningful indication of COVID-19 resolution. Furthermore, Module B contains proteins in the “cholesterol metabolism” pathway (Figure 2F and Table 1), which participate in high-density lipoprotein (HDL) cholesterol metabolism ^43,44^. These data are consistent with a report of low HDL cholesterol levels in patients with severe COVID-19 ^45^. Taken together, the longitudinal increase in protein sets in Module B during disease resolution point to key organ and cellular functions required to reestablish tissue homeostasis and a disease-free state.

### Convalescent plasma transfusion is associated with the disappearance of dysfunctional monocytes

We next utilized single-cell RNA sequencing to analyze transcriptional changes occurring in peripheral blood cells collected from the patient on the day of (prior to) the first transfusion with her relative’s plasma (i.e., day 54 post-symptom onset), and multiple times thereafter (see Fig. 1A for blood collection points). Unsupervised clustering identified one data cluster that was observed pre-transfusion but was absent at subsequent days (Figure 3A). Cell type identification analysis using the Seurat package ^46^ identified this data cluster as being constituted by monocytes (Figure 3B). We next performed Hallmark gene set enrichment analysis to identify genes that best distinguished the monocyte cluster unique to the pre-transfusion state from monocytes in other data clusters. The top hits among upregulated genes [positive normalized enrichment score (NES)] included genes involved in responses to interferon (IFN) alpha (type I), IFN gamma (type II), inflammatory responses, and complement and coagulation (Figure 3C). While type I IFNs are key mediators of antiviral responses ^47^, their excessive activation can exacerbate hyperinflammation and the associated severe manifestations of COVID-19 ^48^. Similar considerations can be made for type II IFN, which may be important for antiviral defense but may worsen systemic inflammation and increase organ damage when produced at persistently high levels ^47^. Thus, the hyperactivation of IFN-γ observed in this patient may have contributed to hyperinflammation and COVID-19 severity. The top hits among downregulated genes (negative NES) comprised MYC targets and signaling by cytokines such as TGFβ and TNF-α (Figure 3C). regulated genes in predicted T cells, monocytes, and NK cells. The color code of the “days post-transfusion” bar corresponds to the color code in panel A.

**Figure 3.**
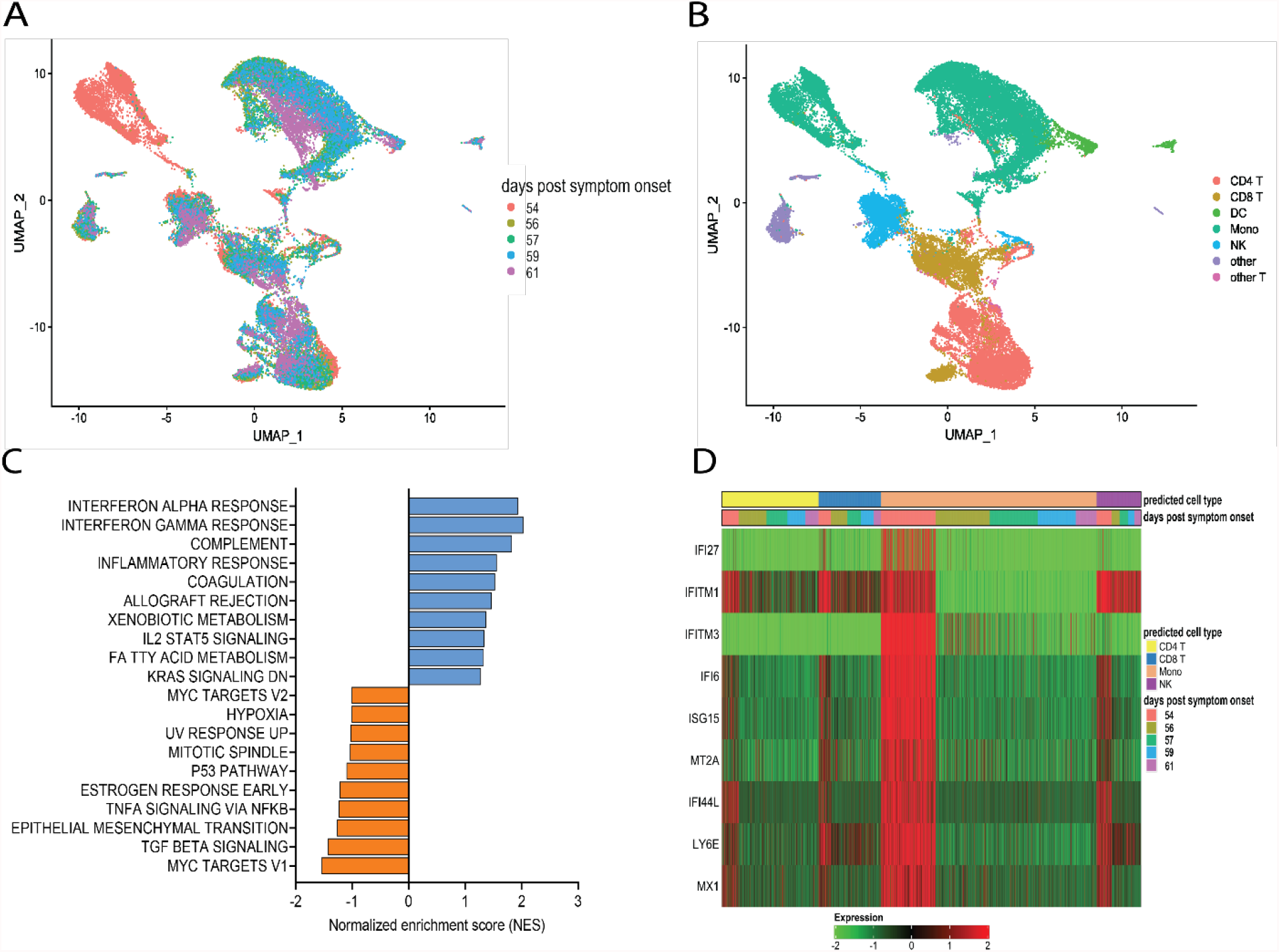

The downregulation of MYC targets is difficult to interpret given the highly diverse cellular functions of the ∼ 400 genes in this gene set. The finding that TNF-α signaling is decreased in the monocyte cluster unique to the pre-transfusion state leads to two sets of considerations. First, the pre-transfusion high levels of TNF-α in plasma (data included in Figure 2A) were likely produced by cell types other than this unique monocyte cluster. Second, an imbalance between signaling by type I IFN (increased) and TNF-α (decreased) in monocytic cells, as observed in this patient, has been proposed to contribute to loss of protective innate immune functions in hyperinflammatory or autoimmune conditions ^49^ and has been described in severe COVID-19 ^50^. Of note, upregulation of both type I IFN and TNF-α responses ^51^ and imbalances in the opposite direction (dampened type I IFN and elevated TNF-α signaling) ^52,53^ have also been reported in severe COVID-19, suggesting that immune dysregulation phenotypes may vary in different patients, presumably depending on context. Furthermore, when we analyzed individual Interferon stimulated gene (ISG) expression, we observed that typical ISGs, such as IFI6, ISG15, IFI44L, and LY6E were most prominently elevated in monocytes prior to transfusion but were also increased in NK cells and T cell subsets, and that this strong IFN response was dampened post-transfusion also in these cell types (Figure 3D). Together, these data show that the beneficial effects of the relative’s plasma transfusion were accompanied by the elimination of a unique population of disease-associated, hyperinflammatory monocytes.

## Discussion

The present case report constitutes, to our knowledge, the first longitudinal molecular and cellular in-depth analysis of COVID-19 resolution. Severe COVID-19 in a woman in her fifties with multiple autoimmune syndrome was associated with SARS-CoV-2 RNAemia, high levels of pro-inflammatory mediators and other potentially harmful plasma factors, dysregulation of coagulation cascades, and abnormally low levels of proteins related to systemic and tissue homeostasis. Following the beneficial convalescent plasma transfusion, the plasma protein landscape was characterized by temporal clusters of cytokines and other plasma proteins, indicative of remarkably dynamic trajectories toward disease resolution. Disease resolution was also associated with post-transfusion disappearance of monocytes characterized by hyperactivation of type I and type II IFN cascades and dampening of TNF-α signaling. It is thus plausible that, by neutralizing virus, the beneficial convalescent plasma caused a drop of viral load in respiratory fluids, effectively dampening IFN responses and the downstream noxious effects of IFN hyperactivation ^48^. These include dysregulation of complement and coagulation functions, potentially leading to lung damage and respiratory failure ^54^. Lung damage might be further compounded by dysfunctional ECM receptors and focal adhesion pathways, as revealed by plasma proteomics, which may underlie the lung injury seen in severe and fatal cases of COVID-19 ^42^. Plasma factors and immune cell types partaking in the changes observed post-transfusion likely identify cellular mechanisms critically associated with disease resolution.

In conclusion, our case report (i) strongly supports the therapeutic use of convalescent plasma in COVID-19 patients unable to mount humoral responses to SARS-CoV-2 infection; (ii) highlights the importance of donor plasma characterization in assessing convalescent plasma therapy efficacy by determining levels, subtype, and specificity of the neutralizing antibodies in the donor plasma; and (iii) identifies biological processes associated with COVID-19 resolution by characterizing their temporal dynamics post-transfusion. These processes may be targets of therapeutic interventions against severe COVID-19, even beyond convalescent plasma therapy.

Our case study has limitations. One is the study of a single patient. The particular characteristics of the case patient, including the underlying immune dysfunction, undoubtedly played a role in both the clinical outcome of the convalescent plasma donation and the ways in which disease resolution occurred. However, most of our findings are corroborated by the rapidly evolving knowledge of the pathways associated with COVID-19 disease and its severity. Thus, we are reasonably confident that our findings are applicable beyond the particular patient under study. Moreover, our report of COVID-19 resolution in a patient unable to generate humoral responses to SARS-CoV-2 infection, together with a previous report of convalescent plasma-associated resolution of COVID-19 in a patient with humoral immunodeficiency ^18^, strongly supports the use of well-characterized convalescent plasma for therapeutic use in COVID-19 patients who are immunocompromised due to underlying defects or immunosuppressive therapies. A second limitation of our work lies in our inability -- despite multiple attempts -- to access the plasma obtained from the first, anonymous donor, which had no beneficial effect on the recipient’s disease. A comparison between the two sets of convalescent plasma samples might have shed light on key therapeutic properties of convalescent plasma for COVID-19. Thirdly, we do not know which organ/cell(s) the viral transcripts we detected in plasma are derived from. Care of the patient required no organ biopsies, and we found no viral RNA in peripheral blood cells (not shown). Nevertheless, our data strongly point to RNAemia as a proxy for severe infection, as also proposed by others ^22^, and perhaps as a tool to monitor viral clearance systemically to identify cases of post-acute viral persistence, which may potentially lead to protracted symptoms ^55^.

## Materials and Methods

### Subject Recruitment

All study activities were approved by the Rutgers Institutional Review Board (Pro2020000655). Informed consent was obtained from the patient in her 50’s (51-55 age range, Caucasian, female) and a relative in their 46-50 age range (Caucasian, male). Blood samples were collected from the two participants at times indicated in the text and in Figure 1A.

### Antibody binding by enzyme-linked immunosorbent assay (ELISA)

Antibody binding was performed by ELISA utilizing SARS-CoV-2 receptor binding domain (RBD) of the Spike protein as solid-phase antigen, as described ^56^. HRP-conjugated mouse anti-human IgG1, IgG2, IgG3, and IgG4 (Southern Biotech, Birmingham, AL, USA) secondary antibodies were used at 1:2,000 dilution. Each sample was tested in duplicate. End-point titers were calculated using background-subtracted data and an established cut-off ^56^.

### Absorption of convalescent plasma with SARS-CoV-2 antigens

96-well ELISA plates (Nunc MaxiSorp, Thermofisher, Rochester, NY, USA) were coated with 500 ng/well of SARS-CoV-2 RBD or N protein at 4°C overnight. Coated plates were washed three times with washing buffer (PBS containing 0.05% Tween 20) (Sigma-Aldrich, St. Louis, MO, USA) blocked with PBS containing 1% BSA (Sigma-Aldrich, St. Louis, MO, USA) for 30 min at 37°C. After washing, plasma samples were diluted 1:10 in PBS containing 1% BSA (Sigma-Aldrich, St. Louis, MO, USA) and incubated overnight at 4°C. Absorption was repeated at least four times utilizing fresh antigen-coated plates at each cycle. To monitor depletion of antigen-specific antibodies prior to use in neutralization assays, antigen-specific IgG titers of untreated and absorbed samples were determined as described above.

### Cell lines

Vero E6 were obtained from the American Type Culture Collection (ATCC) (Manassas, VA, USA); HeLa cells stably expressing ACE2 (HeLa-ACE2) were obtained from Dennis Burton at the Scripps Research Institute ^57^. All cell lines were maintained in high-glucose Dulbecco’s modified Eagle’s medium (DMEM; Corning, Corning, NY, USA) supplemented with 10% fetal bovine serum (FBS; Seradigm, Radnor, PA, USA), 2mM L-glutamine, and 1% penicillin/streptomycin (Corning, Corning, NY, USA), and incubated in humidified atmospheric air containing 5% CO2 at 37°C.

### SARS-CoV-2 virus

The virus stock of mNeonGreen (mNG) SARS-CoV-2 was obtained from Pei-Yong Shi at the University of Texas Medical Branch at Galveston. The virus stock was produced using the virus isolate of the first patient diagnosed in the USA, in which the ORF7 of the viral genome was replaced with the reporter mNG gene ^58^. Propagation of viral stocks was performed with Vero E6 cells using DMEM supplemented with 2% FBS. The virus titers were determined by standard plaque assay utilizing Vero E6 cells and recorded as plaque forming units per milliliter (PFU/mL).

### SARS-CoV-2 neutralization assay

HeLa-Ace2 cells were seeded in 96-well black optical-bottom plates at a density of 1 × 10^4^ cells/well in FluoroBrite DMEM (Thermo Fisher Scientific, Waltham, MA, USA) containing 4% FBS (Seradigm, Radnor, PA, USA), 2mM L-glutamine, and 1% penicillin/streptomycin (Corning, NY, USA), and incubated overnight at 37°C with 5% CO2. On the following day, each sample was subjected to two-fold serial dilution in DMEM without FBS, and incubated with mNG SARS-CoV-2 at 37°C for 1.5 hrs. The virus-plasma mixture was transferred to 96-well plates containing Hela-Ace2 cells at a final multiplicity of infection (MOI) of 0.25 (viral PFU:cell). For each sample, the starting dilution was 1:20 and the final dilution of 1:10,240. After incubating infected cells at 37°C for 20 hrs, mNG SARS-CoV-2 fluorescence was measured using a Cytation™ 5 reader (BioTek, Winooski, VT, USA). Each sample was tested in duplicate. Relative fluorescent units were converted to percent neutralization by normalizing the sample-treatment to non-sample-treatment controls and plotting the data with a nonlinear regression curve fit to determine the titer neutralizing 50% of SARS-CoV-2 fluorescence (NT50).

### Buffy coat isolation and storage

All buffy coats were extracted on the day of receipt at the research laboratory. Briefly, blood tube was centrifuged at 800 × g for 15 mins and plasma was carefully removed, aliquoted, and stored at −80°C. Buffy coat layer was collected, and red blood cells (RBCs) were lysed by incubation in 1× BD Pharm Lyse lysing solution (BD Biosciences, San Jose, CA, USA) at room temperature for 10 mins. Cells were washed 3 times by centrifugation with PBS and counted using a hemocytometer. Buffy coats were cryopreserved in liquid nitrogen in FBS containing 10% dimethyl sulfoxide (DMSO, Thermo Fisher Scientific, Waltham, MA, USA) and stored until use.

### Plasma RT-PCR

Briefly, genomic RNA was extracted from plasma using the RNeasy plus mini kit (QIAGEN, Hilden, Germany). SARS-CoV-2 RNA for nucleocapsid (N) and RNA-dependent RNA polymerase (RdRp) genes was detected by RT-PCR assay, and the results were presented as cycle threshold (Ct).

### Cytokines

Cytokine analysis was performed by the Immune Monitoring and Advanced Genomics Core Facility at Rutgers University using a Luminex® Discovery Assay (R&D System Inc., Minneapolis, MN, USA). The Human Premixed Multi-Analyte Kits were used to detect 48 cytokine/chemokines (CD40L, EGF, Eotaxin, FGF-2, LT-3L, Fractalkine, G-CSF, GROa, IFNα2, IFNγ, IL-1α, IL-1β, IL-1RA, IL-2, IL3, IL-4, IL-5, IL-6, IL-7, IL8, IL-9, IL-10, IL-12 (p40), IL-12 (p70), IL-13, IL-15, IL17A, IL-17E, IL-17F, IL-18, IL-22, IL-27, IP-10, MCP-1, MCP3, M-CSF, MDC, MIG, MIP-1α, MIP-1β, PDGF-AA, PDGF-AB, RANTES, TGFα, TNFα, TNFβ, VEGF-A) according to the manufacturer’s recommendations. Assays were performed in microtiter plates and plates were read with the Luminex 200 instrument. Each sample was measured in duplicate.

### Proteomics

Proteomic analysis was performed by the Rutgers Biological Mass Spectrometry Facility. Plasma samples were prepared for mass spectrometry proteome analysis before or after immunodepletion of abundant plasma proteins. Abundant proteins were removed from 10µl samples using an Agilent Human 14 Multiple Affinity Removal Spin (MARS) Cartridge (Agilent, Santa Clara, CA, USA) using manufacturer’s methods. Protein microchemistry, mass spectrometry and data processing were conducted as described previously ^59^. In brief, protein concentrations of each sample were determined and equal amounts of each (before and after immunodepletion) were proteolytically digested using both filter aided sample preparation ^60^ (FASP) or in-gel protocols as described previously ^59,61^; thus four independent analyses were conducted. Peptides were labeled with TMT 11plex isobaric reagents (Thermo Fisher Scientific, Waltham, MA, USA), pooled then prefractionated by alkaline reverse phase HPLC ^59^. Reporter ions in individual fractions were measured using synchronous precursor selection MS3 methods on a Thermo Eclipse Tribrid mass spectrometer (Thermo Fisher Scientific, Waltham, MA, USA). Peak lists were generated using Proteome Discoverer 2.2 and data were searched using a local implementation of the Global Proteome Machine ^62^. Reporter ion intensities were extracted using in-house scripts (https://github.com/cgermain/IDEAA). Data were normalized to the total reporter ion intensity per channel for each of the four independent analyses to account for differences in protein amounts or labeling efficiency for each sample. Reporter ion intensities were measured after spectra were filtered to remove spectra corresponding to peptides that are non-tryptic or not fully digested, peptides that are incompletely labeled or labeled at positions other than n-termini and lysine, or peptides that contain post-translational modifications that introduce variability

### Multivariate analysis

Plasma cytokine and protein expression measurements were first pre-processed and standardized in MATLAB 2020b (Mathworks; Natick, MA) before subsequent hierarchical clustering and principal component analyses. From the initial set of 48 cytokines and chemokines in the Human Premixed Multi-Analyte Kit, 18 cytokines and chemokines were removed due to poor data quality, where the analyte either fell below the limit of detection or were otherwise unmeasurable in at least one of the samples (EGF, IFNα2, IFNγ, IL-2, IL-3, IL-4, IL-7, IL-10, IL-13, M-CSF, RANTES, TNFβ). Remaining cytokine measurements were log10-transformed and standardized by Z-score. Hierarchical clustering analysis and principal component analysis were performed in MATLAB on the standardized cytokine and protein expression measurements using custom scripts.

### Single cell RNA sequencing

Cells were washed in complete RPMI medium (supplemented with 10% FBS, 2mM L-glutamine, and 1% penicillin/streptomycin), counted, and assessed for viability using a Countess II automated cell counter (Invitrogen, Waltham, MA, USA). Cells were then suspended to 1 × 10^6^/ml in complete RPMI for single-cell emulsion preparation. scRNA-seq GEX libraries were prepared by the Immune Monitoring and Advanced Genomics Rutgers Core Facility according to 10X Genomics specifications. Independent cell suspensions were loaded for droplet-encapsulation by the Chromium Controller (10X Genomics). Single-cell cDNA synthesis, amplification, and sequencing libraries were generated using the Single Cell 5′ Reagent kit (10X Genomics, Pleasanton, CA, USA) following the manufacturer’s instructions. The libraries were shipped to Novogene and sequenced with the Illumina NovaSeq 6000 platform (200 M, 150 bp paired-end reads).

Raw sequencing data were processed using the CellRanger software (version 3.1.0). Reads were aligned to a custom reference genome created with the reference human genome (GRCh38) and SARS-CoV-2 reference genome (NC_045512.2). The resulting unique molecular identifier (UMI) count matrices were imported into R (version 4.0.2) and processed with the R package Seurat (version 4.0.0) ^63^. The data was normalized using SCTransform ^64^ and then dimensionality reduced by finding the top 50 principal components and then using these principal components to run UMAP ^65^. Cell types were identified using Seurat’s MapQuery function, along with the PBMC scRNA-seq datasets ^63^. To account for overclustering, most likely due to variable TCR gene expression in the T cell populations, all TCR genes were removed, the analysis and mapping were repeated, and more homogenous cell populations obtained after mapping. Differential expression results were similar with and without the TCR genes. Doublet analyses were performed using R package Scrublet ^66^. MAST package (version 1.8.2) ^67^ were to identify differential expressing genes across different cell populations. Differentially expressing genes were used with the fast pre-ranked gene set enrichment analysis (fGSEA) package in R for the hallmark pathways ^68^.

## Data Availability

All data produced in the present study are available upon reasonable request to the authors

## Acknowledgements

We thank the COVID-19 patient who prompted this study; Dennis Burton and Pei-Yong Shi for providing biological reagents; David Sleat and Peter Lobel for the plasma proteomic analyses. This work was funded by NIH grants R01 HL149450, R01 HL149450-S1, R61 HD105619, R01 AI158911, U01 AI122285-S1, UL1 TR003017, and R00 GM118907. Work performed at the Rutgers Biological Mass Spectrometry Facility was partially supported by NIH S10 OD025140.

## Author contributions

Conceptualization, V.G., R.U., S.L., M.L.G and N.B.; Methodology, R.D and S.T.; Formal analysis, J.Y., R.D. and N.B.; Investigation, R.U., V.G., R.D., J.V., N.B. and P.K.M.; Resources, A.O., D.H., M.C, S.G., M.P., C.P; A.P. and S.L.; Data Curation, J.V., N.B., V.G., R.U., R.D., S.T., J.Y. and M.L.G; Writing – Original Draft, N.B. and M.L.G; Writing – Review and Editing, M.L.G, N.B., V.G., R.U., R.D., J.Y., S.G., M.P., S.L., and C.P., Visualization, N.B., R.U., V.G., R.D. and J.Y.; Supervision, M.L.G. and S.L.; Funding, J.Y., M.L.G.

## Author declaration

We confirm that all relevant ethical guidelines have been followed and all study activities were approved by the Rutgers Institutional Review Board (Pro2020000655). We also confirm that written consent to publish the present work was obtained from the patient. The signed consent is available for review by the editorial team.

## Notes

### Competing Interest Statement

Rutgers University has filed for patent protection for various aspects of anti-SARS-CoV-2 antibody detection and its uses. M.L.G. is a consultant for Biomerieux.

### Author Declarations

All study activities were approved by the Rutgers Institutional Review Board (Pro2020000655).

## References

1. Bakhiet M, Taurin S. SARS-CoV-2: Targeted managements and vaccine development. Cytokine Growth Factor Rev. 2021;58:16–29.

2. Nabil A, Uto K, Elshemy MM, et al. Current coronavirus (SARS-CoV-2) epidemiological, diagnostic and therapeutic approaches: An updated review until June 2020. Excli j. 2020;19:992–1016.

3. Pavan M, Bolcato G, Bassani D, Sturlese M, Moro S. Supervised Molecular Dynamics (SuMD) Insights into the mechanism of action of SARS-CoV-2 main protease inhibitor PF-07321332. J Enzyme Inhib Med Chem. 2021;36(1):1646–1650.

4. Fischer W, Eron JJ, Holman W, et al. Molnupiravir, an Oral Antiviral Treatment for COVID-19. medRxiv. 2021.

5. https://www.merck.com/news/merck-and-ridgeback-biotherapeutics-provide-update-on-results-from-move-out-study-of-molnupiravir-an-investigational-oral-antiviral-medicine-in-at-risk-adults-with-mild-to-moderate-covid-19/.

6. https://investor.regeneron.com/static-files/969bdb0b-53f5-46c7-94fb-7473ee7f5be3.

7. Duan K, Liu B, Li C, et al. Effectiveness of convalescent plasma therapy in severe COVID-19 patients. Proc Natl Acad Sci U S A. 2020;117(17):9490–9496.

8. Liu STH, Lin HM, Baine I, et al. Convalescent plasma treatment of severe COVID-19: a propensity score-matched control study. Nat Med. 2020;26(11):1708–1713.

9. Shen C, Wang Z, Zhao F, et al. Treatment of 5 Critically Ill Patients With COVID-19 With Convalescent Plasma. Jama. 2020;323(16):1582–1589.

10. Zhang B, Liu S, Tan T, et al. Treatment With Convalescent Plasma for Critically Ill Patients With Severe Acute Respiratory Syndrome Coronavirus 2 Infection. Chest. 2020;158(1):e9–e13.

11. Lucas C, Klein J, Sundaram M, et al. Kinetics of antibody responses dictate COVID-19 outcome. medRxiv. 2020.

12. https://www.who.int/publications/i/item/WHO-2019-nCoV-therapeutics-2021.4.

13. Simonovich VA, Burgos Pratx LD, Scibona P, et al. A Randomized Trial of Convalescent Plasma in Covid-19 Severe Pneumonia. N Engl J Med. 2021;384(7):619–629.

14. Agarwal A, Mukherjee A, Kumar G, Chatterjee P, Bhatnagar T, Malhotra P. Convalescent plasma in the management of moderate covid-19 in adults in India: open label phase II multicentre randomised controlled trial (PLACID Trial). Bmj. 2020;371:m3939.

15. Jones JM, Faruqi AJ, Sullivan JK, Calabrese C, Calabrese LH. COVID-19 Outcomes in Patients Undergoing B Cell Depletion Therapy and Those with Humoral Immunodeficiency States: A Scoping Review. Pathog Immun. 2021;6(1):76–103.

16. Shanehbandi D, Majidi J, Kazemi T, Baradaran B, Aghebati-Maleki L. CD20-based Immunotherapy of B-cell Derived Hematologic Malignancies. Curr Cancer Drug Targets. 2017;17(5):423–444.

17. Apostolidis SA, Kakara M, Painter MM, et al. Cellular and humoral immune responses following SARS-CoV-2 mRNA vaccination in patients with multiple sclerosis on anti-CD20 therapy. Nat Med. 2021.

18. Honjo K, Russell RM, Li R, et al. Convalescent plasma-mediated resolution of COVID-19 in a patient with humoral immunodeficiency. Cell Rep Med. 2021;2(1):100164.

19. Böhm I. Decrease of B-cells and autoantibodies after low-dose methotrexate. Biomed Pharmacother. 2003;57(7):278–281.

20. Bonelli MM, Mrak D, Perkmann T, Haslacher H, Aletaha D. SARS-CoV-2 vaccination in rituximab-treated patients: evidence for impaired humoral but inducible cellular immune response. Ann Rheum Dis. 2021;80(10):1355–1356.

21. Finkel Y, Mizrahi O, Nachshon A, et al. The coding capacity of SARS-CoV-2. Nature. 2021;589(7840):125–130.

22. Gutmann C, Takov K, Burnap SA, et al. SARS-CoV-2 RNAemia and proteomic trajectories inform prognostication in COVID-19 patients admitted to intensive care. Nat Commun. 2021;12(1):3406.

23. Prebensen C, Myhre PL, Jonassen C, et al. Severe Acute Respiratory Syndrome Coronavirus 2 RNA in Plasma Is Associated With Intensive Care Unit Admission and Mortality in Patients Hospitalized With Coronavirus Disease 2019. Clin Infect Dis. 2021;73(3):e799–e802.

24. Lucas C, Wong P, Klein J, et al. Longitudinal analyses reveal immunological misfiring in severe COVID-19. Nature. 2020;584(7821):463–469.

25. Kuleshov MV, Jones MR, Rouillard AD, et al. Enrichr: a comprehensive gene set enrichment analysis web server 2016 update. Nucleic Acids Res. 2016;44(W1):W90–97.

26. Ramlall V, Thangaraj PM, Meydan C, et al. Immune complement and coagulation dysfunction in adverse outcomes of SARS-CoV-2 infection. Nat Med. 2020;26(10):1609–1615.

27. Lo MW, Kemper C, Woodruff TM. COVID-19: Complement, Coagulation, and Collateral Damage. J Immunol. 2020;205(6):1488–1495.

28. Ricklin D, Reis ES, Lambris JD. Complement in disease: a defence system turning offensive. Nat Rev Nephrol. 2016;12(7):383–401.

29. Semeraro N, Colucci M. The Prothrombotic State Associated with SARS-CoV-2 Infection: Pathophysiological Aspects. Mediterr J Hematol Infect Dis. 2021;13(1):e2021045.

30. Gómez-Rial J, Currás-Tuala MJ, Rivero-Calle I, et al. Increased Serum Levels of sCD14 and sCD163 Indicate a Preponderant Role for Monocytes in COVID-19 Immunopathology. Front Immunol. 2020;11:560381.

31. Zhu P, Lv C, Fang C, et al. Heat Shock Protein Member 8 Is an Attachment Factor for Infectious Bronchitis Virus. Front Microbiol. 2020;11:1630.

32. Taguwa S, Yeh MT, Rainbolt TK, et al. Zika Virus Dependence on Host Hsp70 Provides a Protective Strategy against Infection and Disease. Cell Rep. 2019;26(4):906–920.e903.

33. Ramos CHI, Ayinde KS. Are Hsp90 inhibitors good candidates against Covid-19? Curr Protein Pept Sci. 2020.

34. Paladino L, Vitale AM, Caruso Bavisotto C, et al. The Role of Molecular Chaperones in Virus Infection and Implications for Understanding and Treating COVID-19. J Clin Med. 2020;9(11).

35. Murdaca G, Contini P, Cagnati P, et al. Behavior of soluble HLA-A, -B, -C and HLA-G molecules in patients with chronic hepatitis C virus infection undergoing pegylated interferon-α and ribavirin treatment: potential role as markers of response to antiviral therapy. Clin Exp Med. 2017;17(1):93–100.

36. Cooper EH, Forbes MA, Hambling MH. Serum beta 2-microglobulin and C reactive protein concentrations in viral infections. J Clin Pathol. 1984;37(10):1140–1143.

37. Conca W, Alabdely M, Albaiz F, et al. Serum β2-microglobulin levels in Coronavirus disease 2019 (Covid-19): Another prognosticator of disease severity? PLoS One. 2021;16(3):e0247758.

38. Owusu BY, Zimmerman KA, Murphy-Ullrich JE. The role of the endoplasmic reticulum protein calreticulin in mediating TGF-β-stimulated extracellular matrix production in fibrotic disease. J Cell Commun Signal. 2018;12(1):289–299.

39. EchavarrÍa-Consuegra L, Cook GM, Busnadiego I, et al. Manipulation of the unfolded protein response: A pharmacological strategy against coronavirus infection. PLoS Pathog. 2021;17(6):e1009644.

40. van de Veerdonk FL, Netea MG, van Deuren M, et al. Kallikrein-kinin blockade in patients with COVID-19 to prevent acute respiratory distress syndrome. Elife. 2020;9.

41. Bonnans C, Chou J, Werb Z. Remodelling the extracellular matrix in development and disease. Nat Rev Mol Cell Biol. 2014;15(12):786–801.

42. Leng L, Cao R, Ma J, et al. Pathological features of COVID-19-associated lung injury: a preliminary proteomics report based on clinical samples. Signal Transduct Target Ther. 2020;5(1):240.

43. Actis Dato V, Chiabrando GA. The Role of Low-Density Lipoprotein Receptor-Related Protein 1 in Lipid Metabolism, Glucose Homeostasis and Inflammation. Int J Mol Sci. 2018;19(6).

44. Mantuano E, Brifault C, Lam MS, Azmoon P, Gilder AS, Gonias SL. LDL receptor-related protein-1 regulates NFκB and microRNA-155 in macrophages to control the inflammatory response. Proc Natl Acad Sci U S A. 2016;113(5):1369–1374.

45. Begue F, Tanaka S, Mouktadi Z, et al. Altered high-density lipoprotein composition and functions during severe COVID-19. Sci Rep. 2021;11(1):2291.

46. Stuart T, Butler A, Hoffman P, et al. Comprehensive Integration of Single-Cell Data. Cell. 2019;177(7):1888–1902.e1821.

47. Lee AJ, Ashkar AA. The Dual Nature of Type I and Type II Interferons. Front Immunol. 2018;9:2061.

48. Lee JS, Shin EC. The type I interferon response in COVID-19: implications for treatment. Nat Rev Immunol. 2020;20(10):585–586.

49. Cantaert T, Baeten D, Tak PP, van Baarsen LG. Type I IFN and TNFα cross-regulation in immune-mediated inflammatory disease: basic concepts and clinical relevance. Arthritis Res Ther. 2010;12(5):219.

50. Wilk AJ, Lee MJ, Wei B, et al. Multi-omic profiling reveals widespread dysregulation of innate immunity and hematopoiesis in COVID-19. J Exp Med. 2021;218(8).

51. Lee JS, Park S, Jeong HW, et al. Immunophenotyping of COVID-19 and influenza highlights the role of type I interferons in development of severe COVID-19. Sci Immunol. 2020;5(49).

52. Hadjadj J, Yatim N, Barnabei L, et al. Impaired type I interferon activity and inflammatory responses in severe COVID-19 patients. Science. 2020;369(6504):718–724.

53. Blanco-Melo D, Nilsson-Payant BE, Liu WC, et al. Imbalanced Host Response to SARS-CoV-2 Drives Development of COVID-19. Cell. 2020;181(5):1036–1045.e1039.

54. Frantzeskaki F, Armaganidis A, Orfanos SE. Immunothrombosis in Acute Respiratory Distress Syndrome: Cross Talks between Inflammation and Coagulation. Respiration. 2017;93(3):212–225.

55. Huang Z, Ning B, Yang HS, et al. Sensitive tracking of circulating viral RNA through all stages of SARS-CoV-2 infection. J Clin Invest. 2021;131(7).

56. Datta P, Ukey R, Bruiners N, et al. Highly versatile antibody binding assay for the detection of SARS-CoV-2 infection and vaccination. J Immunol Methods. 2021;499:113165.

57. Rogers TF, Zhao F, Huang D, et al. Isolation of potent SARS-CoV-2 neutralizing antibodies and protection from disease in a small animal model. Science. 2020;369(6506):956–963.

58. Xie X, Muruato A, Lokugamage KG, et al. An Infectious cDNA Clone of SARS-CoV-2. Cell Host Microbe. 2020;27(5):841–848.e843.

59. Tannous A, Boonen M, Zheng H, et al. Comparative Analysis of Quantitative Mass Spectrometric Methods for Subcellular Proteomics. J Proteome Res. 2020;19(4):1718–1730.

60. Wiśniewski JR. Quantitative Evaluation of Filter Aided Sample Preparation (FASP) and Multienzyme Digestion FASP Protocols. Anal Chem. 2016;88(10):5438–5443.

61. Sleat DE, Della Valle MC, Zheng H, Moore DF, Lobel P. The mannose 6-phosphate glycoprotein proteome. J Proteome Res. 2008;7(7):3010–3021.

62. Beavis RC. Using the global proteome machine for protein identification. Methods Mol Biol. 2006;328:217–228.

63. Hao Y, Hao S, Andersen-Nissen E, et al. Integrated analysis of multimodal single-cell data. Cell. 2021;184(13):3573–3587.e3529.

64. Hafemeister C, Satija R. Normalization and variance stabilization of single-cell RNA-seq data using regularized negative binomial regression. Genome Biol. 2019;20(1):296.

65. Becht E, McInnes L, Healy J, et al. Dimensionality reduction for visualizing single-cell data using UMAP. Nat Biotechnol. 2018.

66. Wolock SL, Lopez R, Klein AM. Scrublet: Computational Identification of Cell Doublets in Single-Cell Transcriptomic Data. Cell Syst. 2019;8(4):281–291.e289.

67. Finak G, McDavid A, Yajima M, et al. MAST: a flexible statistical framework for assessing transcriptional changes and characterizing heterogeneity in single-cell RNA sequencing data. Genome Biol. 2015;16:278.

68. Liberzon A, Birger C, Thorvaldsdóttir H, Ghandi M, Mesirov JP, Tamayo P. The Molecular Signatures Database (MSigDB) hallmark gene set collection. Cell Syst. 2015;1(6):417–425.

