## Supplemental_figures_legends_tables for "Biologic correlates of beneficial convalescent plasma therapy in a COVID-19 patient reveal disease resolution mechanisms"

### Supplemental file

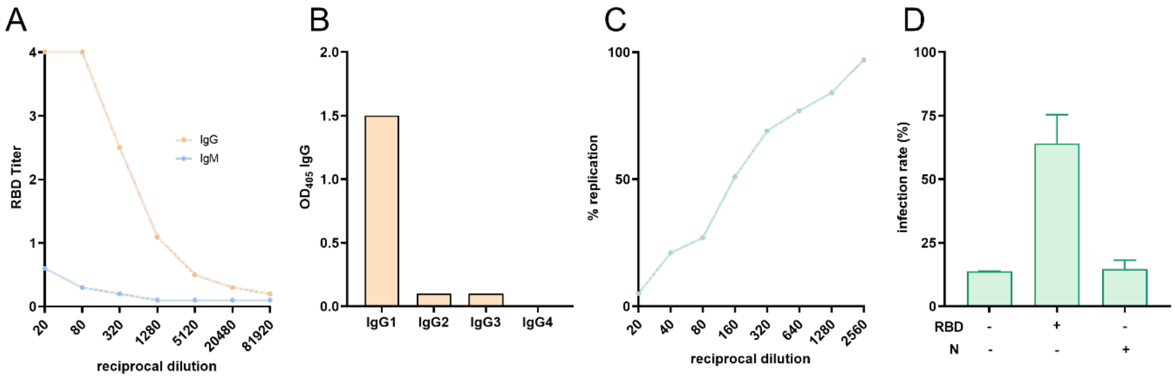

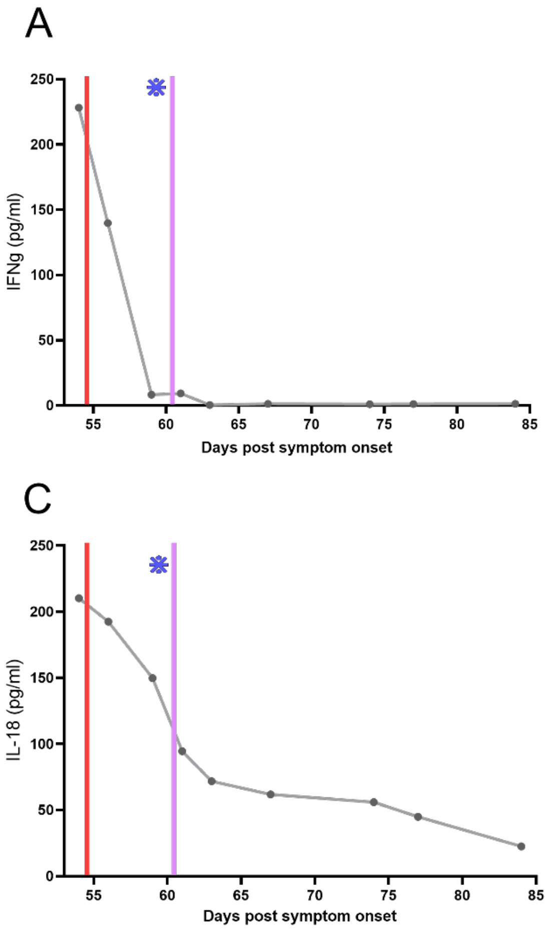
A
IFNg (pg/ml)
Days post symptom onset
B
IL-10 (pg/ml)
Days post symptom onset
C
IL-18 (pg/ml)
Days post symptom onset
D
M-CSF (pg/ml)
Days post symptom onset

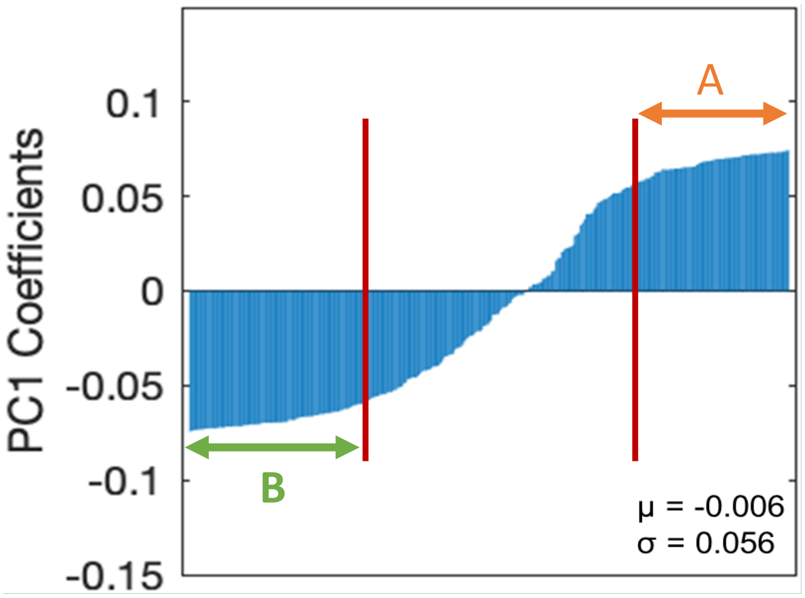


**Supplemental Figure 3.** Coefficients for proteins from Principal Component 1 (PC1) from the proteomic analyses (Figure 2A). Proteins with decreasing expression after convalescent plasma transfusion (Module A, orange arrow) possessed coefficients greater than one standard deviation from the mean, while proteins with increasing expression (Module B, green arrow) possessed coefficients less than one standard deviation from the mean.
